# Nurse-Led Discharge Planning and Its Impact on Psychological Outcomes and Readmissions in Heart Failure: A Literature Review

**DOI:** 10.1101/2025.09.17.25335454

**Authors:** Ida Ayu Agung Laksmi, Moses Glorino Rumambo Pandin, Tintin Sukartini

## Abstract

**Background:** Heart failure (HF) is a leading cause of morbidity, mortality, and hospital readmissions worldwide, creating significant clinical, psychological, and financial burdens. Despite advances in treatment, frequent rehospitalizations and poor psychosocial outcomes remain pressing challenges, partly due to inadequate discharge planning and transitional care.

**Objective:** This literature review aimed to examine the effectiveness of discharge planning—particularly nurse-led interventions—on psychological outcomes and hospital readmissions among patients with HF, drawing on studies published between 2020 and 2025.

**Method:** Guided by the PRISMA framework, a systematic search was conducted in Scopus, ProQuest, EBSCOHost, and Web of Science. Eligible studies included adult HF patients, assessed discharge planning or nurse-led interventions, and reported outcomes related to readmissions, psychological status, self-care, or quality of life. Ten studies met the inclusion criteria, encompassing quasi-experimental, cross-sectional, pilot study, and randomized controlled trial designs

**Results:** Across the reviewed studies, structured and individualized discharge planning was consistently associated with improved self-care behaviors, disease knowledge, treatment adherence, and psychological well-being. Nurse-led education, particularly when delivered through methods such as teach-back, demonstrated significant reductions in anxiety, depression, and caregiver dependency. Several interventions also reported reduced 30-day readmission rates and cost savings, though findings on rehospitalization were mixed due to variations in intervention design, follow-up duration, and health system.

**Conclusion:** Evidence supports discharge planning, especially nurse-led and patient-cantered approaches as an effective strategy to improve self-care, enhance psychological outcomes, and reduce rehospitalizations in HF patients. Future research should prioritize multi-center randomized controlled studies, longer follow-up periods, and integration of digital health tools to strengthen transitional care and sustain improvements over time.

## INTRODUCTION

Heart failure (HF) is one of the most prevalent cardiovascular diseases worldwide, associated with high mortality and morbidity rates and considerable financial burden, particularly among older adults (Zdanowicz 2002; Prihatiningsih & Sudyasih 2018; Saelan, Dzurriyatun Toyyibah 2021). he growing number of patients, frequent hospital readmissions, and rising disability and mortality rates have made HF a major concern in cardiology (Saelan, Dzurriyatun Toyyibah 2021). According to the American Heart Association (AHA, 2019), approximately 15 million people worldwide experience HF symptoms, and this number is projected to increase to 23.6 million by 2030 (Zdanowicz 2002).

Despite advances in pharmacological and device-based therapies, hospital readmission remains a major challenge in HF management, with nearly one in four patients rehospitalized within 30 days of discharge (Kinugasa et al. 2024; Averbuch et al. 2025) . The one-year mortality rate among HF patients in the United States has been reported at 37.5%, with a hospitalization rate of 30.9% (Koikai & Khan 203). These recurrent admissions not only escalate healthcare costs but also compromise patients’ quality of life and psychological well-being (Puspitawati & Widani 2024).

The transition from hospital to home represents a critical period of vulnerability, as patients must adapt to complex treatment regimens, monitor symptoms, and adhere to lifestyle modifications. However, many patients lack the necessary knowledge, skills, or confidence to effectively manage their condition post-discharge. Inadequate discharge planning and poor continuity of care therefore contribute substantially to preventable readmissions and poorer health outcomes (Dhaliwal & Dang 2025).

Structured discharge planning has emerged as a cornerstone of transitional care. When delivered in a comprehensive, individualized, and nurse-led manner, discharge planning has been shown to improve self-care behaviors, strengthen treatment adherence, and enhance psychosocial outcomes, including reductions in anxiety and depression (Gonçalves-Bradley et al. 2022; Faessler et al. 2023). Interventions such as patient education, self-management support, and follow-up care have demonstrated potential in addressing not only clinical but also psychosocial aspects of HF management (Chew et al. 2021; Hany & Vatmasari 2023; Suharsono et al. 2024). Nevertheless, the effectiveness of these strategies varies across settings and populations, highlighting the need to synthesize recent evidence.

Therefore, the problem emergence is that hospital readmissions and psychological distress continue to be major challenges in heart failure care, largely due to inadequate discharge planning and insufficient transitional care, despite advances in available therapies. This literature review aims to examine the effectiveness of discharge planning on psychological outcomes and hospital readmissions among patients with heart failure, focusing on studies published between 2020 and 2025.

## METHOD

This study employed a literature review approach to examine the components that support discharge planning in facilitating the transition of care among patients with heart failure. Guided by the PRISMA (Preferred Reporting Items for Systematic Reviews and Meta-Analyses) framework, a systematic search was conducted across Scopus, ProQuest, EBSCOHost, and Web of Science to identify relevant studies published between 2020 and 2025. The search strategy applied a combination of keywords and Medical Subject Headings (MeSH), including “heart failure,” “discharge planning,” “nurse-led intervention,” “self-care,” “patient education,” and “readmission,” with Boolean operators (AND, OR) used to refine the results.

Reference lists of selected articles were also manually screened to capture additional eligible studies. Studies were included if they focused on adult patients diagnosed with heart failure, examined the impact of discharge planning or nurse-led interventions on clinical, psychosocial, or behavioral outcomes, and reported at least one measurable outcome such as readmission rates, quality of life, psychological status, self-care, or healthcare utilization. Only studies published in English within the specified period were considered. Exclusion criteria included editorials, reviews, conference abstracts, case reports, studies lacking primary data, and interventions not directly related to discharge planning or transitional care. All search results were imported into Mendeley for reference management, and duplicates were removed. Titles and abstracts were independently screened by two reviewers, with full-texts of potentially relevant articles retrieved and assessed for eligibility. Disagreements during the selection process were resolved by consensus. Data extraction was carried out using a standardized form that captured key information including author(s), year of publication, study design and setting, sample size and characteristics, type of intervention, outcome measures, and major findings.

## RESULT

A total of 1,341 records were identified from Scopus (n=241), ProQuest (n=624), EBSCOHost (n=92), and Web of Science (n=384). After duplicate removal, 577 articles remained. Following title and abstract screening, 121 were assessed, of which 32 underwent full-text evaluation. Finally, 10 studies met the inclusion criteria and were included in this review. The PRISMA flow diagram summarizing the screening process is presented in Figure 1.

**Fig 1.**
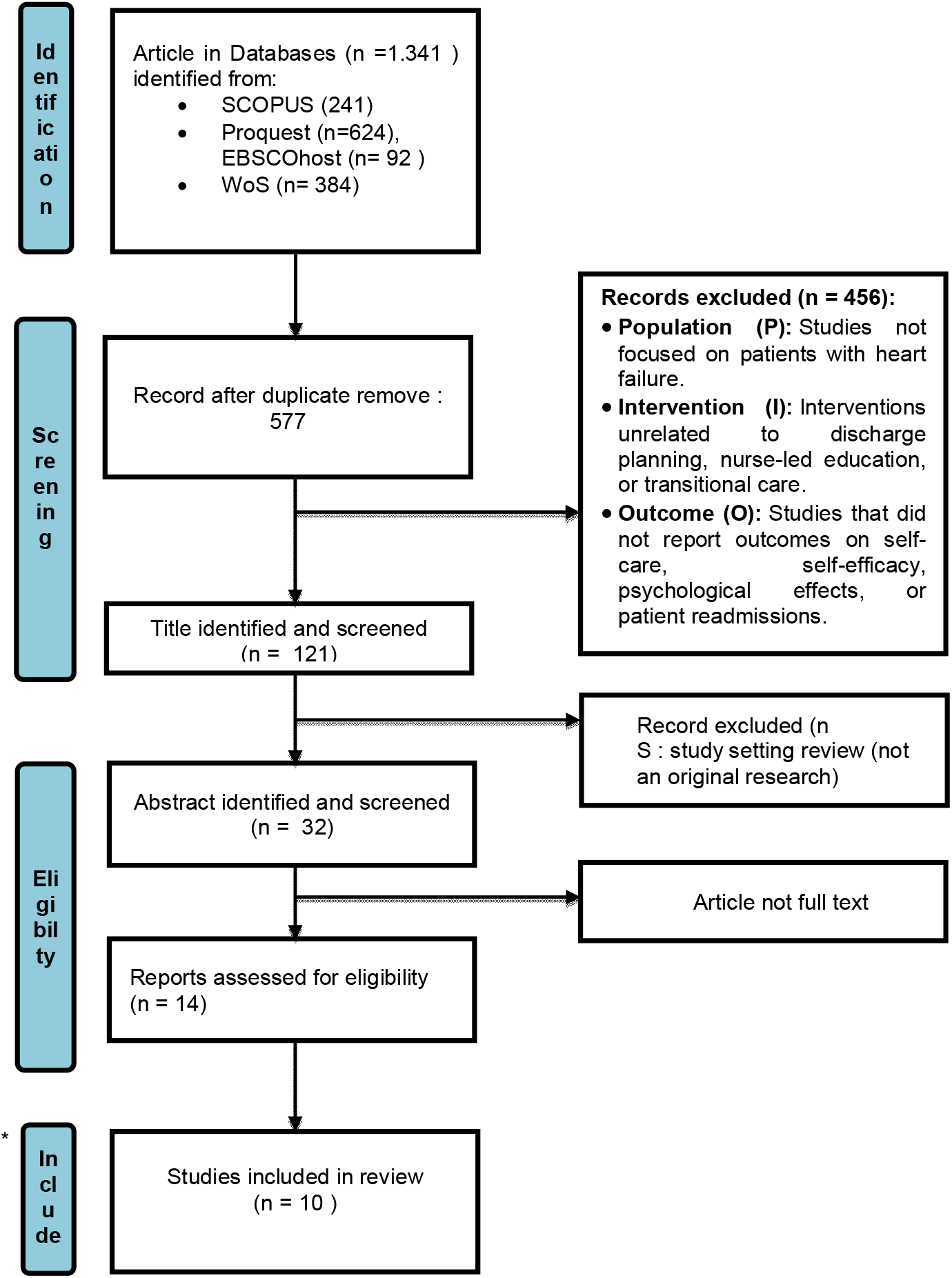
PRISMA Flow-chart.

The 10 studies comprised a mix of randomized controlled trials (RCTs), quasi-experimental designs, cross-sectional analyses, pilot studies, and quality improvement projects, conducted in diverse settings across the United States, Pakistan, Iran, Poland, Taiwan, Hong Kong, Bosnia and Herzegovina, and other regions. Sample sizes ranged from 20 to 1,584 patients. Interventions varied from structured discharge education, nurse-led self-care programs, and teach-back methods, to transitional care models involving follow-up via telephone or digital platforms.

**Table 1.** Result of Article Analysis.

| No | Author(s),<br>Year | Objective | Design &<br>Sample | Intervention | Key Findings |
| --- | --- | --- | --- | --- | --- |
| 1 | Abouzid & Siddiqi, 2024 | Explore relationship between age/gender and discharge disposition in HF | Cross-sectional; 1,584 readmitted HF patients ($\geq 65$ yrs), US | Analysis of discharge planning practices | Effective discharge planning reduced readmissions regardless of age/gender |
| 2 | Hosseini et al., 2023 | Effect of self-management-based discharge planning on anxiety, depression, and readmission | Quasi-experimental; 80 HF patients, Iran | Structured discharge + telephone follow-up (up to 3 months) | Significant ↓ anxiety & depression ($p < 0.001$); no significant difference in readmission rates |
| 3 | Weeks, Kile & Garber, 2020 | Evaluate nurse discharge navigator on 30-day readmissions | Prospective cohort pilot; 28 HF patients, US | Discharge navigator model in community hospital | 30-day readmissions ↓ (Jan 24% → Apr 11%); potential cost avoidance ≈ \$405,316 |
| 4 | Iqbal et al., 2020 | Impact of nursing discharge instructions on post-discharge care | Quasi-experimental; 80 HF patients, Pakistan | Enhanced nurse-led discharge education (vs. routine care) | Improved awareness, adherence, and disease management ($p = 0.001$) |
| 5 | Oh et al., 2023 | Effect of teach-back method on self-care & healthcare utilization | RCT; 100 HF patients, Korea | HEART program® + teach-back vs. usual education | Significant ↑ self-care maintenance ($p = 0.001$), symptom perception ($p < 0.001$), and efficacy |
| 6 | Kolasa et al., 2021 | Assess nurse-led “Weak Heart” education model | Quasi-experimental; 259 HF patients, Poland | Nurse-led structured education | Significant ↑ HF knowledge ($p < 0.001$) and self-care behaviors ($p < 0.001$) at 3 months |
| 7 | Vijaya Kumari et al., 2025 | Evaluate standardized nurse-led education on readmissions | Quality improvement project; 30 HF patients, India | Comprehensive discharge education on self-care & medication | 30-day readmissions ↓ from 33% → 13%; significant ↑ knowledge and adherence |
| 8 | Huang & Chair, 2025 | Develop & test nurse-led self-regulation model | Pilot RCT; 26 HF patients, Hong Kong | Common-sense model of self-regulation-based program | Improved illness perceptions, self-efficacy, and self-care behaviors |
| 9 | Dizdarevic-Hudic et al., 2025 | Evaluate nurse-led patient education on QoL & rehospitalization | RCT; 64 HF rEF patients, Bosnia & Herzegovina | Structured nurse-led HF education vs. routine care | Improved QoL; rehospitalizations 0% (intervention) vs. 34% (control) |
| 10 | Yeh & Tsai, 2024 | Assess transitional care program on depression, self-efficacy, and self-care | RCT; 20 HF patients, Taiwan | Transition care + routine care vs. routine care only | Significant ↑ self-efficacy ($p = 0.011$) and self-care ($p < 0.01$) after 1 month |

## DISCUSSION

This literature consists of 10 articles exploring the effect of nurse led education or discharge -planning in heart failure patients. Based on the analysis of 10 journals, the study used a quantitative method with a quasy experimental, cross-sectional stud, A pilot study methods which examines the use of discharge planning in preparing patient care in hospitals. Data collection was done through observation and medical records. The sample in this study were heart failure patients, who returned to hospital care. To assess compliance in the treatment of discharge planning that has been given by nurses, the distribution of samples came from several countries including the United States, Pakistan, Iran, Poland, Taiwan, Hong Kong, Bosnia and Herzegovina, and other regions.

Effective discharge planning has consistently been shown to play a pivotal role in optimizing outcomes for patients with heart failure (HF). Across diverse settings and methodologies, the reviewed studies emphasize the importance of structured, nurse-led, and patient-centered interventions in reducing readmissions, improving self-care, and enhancing quality of life.

Several studies underscore the central role of discharge education in fostering effective self-management. For example, Iqbal et al. (2020) demonstrated that nurse-delivered discharge instructions significantly improved patients’ awareness, medication adherence, and disease management competencies. Similarly, Oh et al. (2023) confirmed the effectiveness of the teach-back method, showing significant improvements in self-care maintenance, symptom perception, self-care management, and self-care efficacy. These findings are consistent with Kolasa et al. (2021), who reported that a nurse-led educational program improved disease knowledge and self-care behaviors over a three-month period, and with Vijaya Kumari et al. (2025), who observed notable reductions in 30-day readmissions and improvements in self-care adherence following standardized nurse-led education. Taken together, these results highlight patient education as a cornerstone of successful discharge interventions.

Beyond educational strategies, several studies examined the psychological and behavioral impacts of discharge planning. Hosseini et al. (2023) found that self-management-based discharge planning significantly reduced anxiety and depression, while a newly developed transition care program improved self-efficacy and self-care behaviors after one month. These results suggest that discharge interventions have benefits that extend beyond physical outcomes, enhancing psychological well-being and patients’ confidence in managing their illness. The integration of psychosocial support into discharge planning may therefore be essential for long-term success.

The influence of discharge interventions on readmissions and healthcare utilization has been another focal point of research. Abouzid and Siddiqi (2024) highlighted that effective discharge planning reduced readmissions for acute exacerbation of chronic heart failure regardless of age or gender, while Weeks, Kile, and Garber (2020) reported a reduction in heart failure readmissions during the implementation of a nurse discharge navigator, accompanied by substantial cost savings. Similarly, Dizdarevic-Hudic et al. (2025) demonstrated that none of the patients receiving structured nurse-led education were rehospitalized, in contrast to a 34% rehospitalization rate in the control group. However, not all findings were consistent; Hosseini et al. (2023) did not observe significant differences in readmission frequency or duration, suggesting that while educational and psychosocial interventions improve patient-reported outcomes, broader systemic factors may influence rehospitalization rates.

Overall, these studies converge on the conclusion that structured, nurse-led discharge planning is an effective strategy to improve patient outcomes in heart failure. Interventions focusing on education, psychosocial support, and transitional care enhance self-care behaviors, reduce psychological distress, and lower hospital utilization. Nonetheless, gaps remain. Many studies relied on quasi-experimental or small-scale designs, limiting generalizability. The heterogeneity of interventions and follow-up periods also complicates comparisons. Future research should prioritize large-scale randomized controlled trials, explore the integration of digital health and telemonitoring, and investigate strategies to sustain improvements over longer periods.

### Implication

The findings carry important implications for both clinical practice and future research. From a clinical perspective, the consistent improvements in self-care, knowledge, and psychological outcomes indicate that discharge planning should be integrated as a core nursing responsibility in the management of heart failure patients. Structured education, particularly using evidence-based methods such as teach-back, should be standardized as part of discharge protocols. Moreover, interventions that extend support beyond hospitalization—such as telephone follow-up or transition care programs, appear to enhance sustainability and may reduce readmissions more effectively than single-session approaches.

From a health systems perspective, the demonstrated reductions in readmission rates and cost savings (Weeks et al., 2020; Vijaya Kumari et al., 2025) highlight the economic value of investing in nurse-led discharge programs. Policy makers and hospital administrators should consider scaling these interventions to reduce healthcare utilization and financial burden.

For future research, large-scale, multi-center randomized controlled trials with standardized outcome measures are needed to validate the effectiveness of specific components of discharge interventions. Longer follow-up periods would also help determine whether improvements in self-care and psychological well-being translate into sustained reductions in rehospitalization. Additionally, integrating digital health tools, such as telemonitoring and mobile applications, may further strengthen patient engagement and continuity of care, especially for older populations who face challenges with traditional self-management.

### Limitation

First, heterogeneity in interventions including educational content, delivery methods, and follow-up duration—complicates direct comparisons and synthesis across studies. For instance, while some interventions incorporated ongoing telephonic follow-up (Hosseini et al., 2023; Iqbal et al., 2020), others were limited to a single structured educational session (Oh et al., 2023; Vijaya Kumari et al., 2025). Second, outcome measures were not standardized, ranging from self-care behaviors and psychosocial indicators to readmission rates and cost savings. This variability makes it difficult to establish consensus on the most effective intervention components. Third, most studies were conducted within single institutions or community hospitals, raising questions about scalability and applicability across different health care systems, especially in low-resource settings. Finally, this review did not include a meta-analysis, which limits the ability to quantify pooled effect sizes and establish stronger causal inferences.

## CONCLUSION

This review provides robust evidence that structured and nurse-led discharge planning significantly improves outcomes for patients with heart failure. Interventions consistently enhanced self-care behaviors, disease knowledge, treatment adherence, and psychological well-being, including reductions in anxiety and depression. Several studies also demonstrated reductions in 30-day readmission rates and healthcare costs, although findings were not uniformly consistent due to heterogeneity in study design and healthcare settings. Overall, nurse-led, patient-centered discharge planning represents an effective and cost-efficient strategy to optimize transitional care for HF patients. By addressing both clinical and psychosocial needs, these interventions offer a comprehensive approach that extends beyond hospitalization and into sustained self-management.

## Data Availability

All data produced in the present study are available upon reasonable request to the authors

